# Fidaxomicin as first line: What will it cost in the USA and Canada?

**DOI:** 10.1101/2021.11.03.21265881

**Authors:** Devangi Patel, Julien Senecal, Brad Spellberg, Andrew M. Morris, Lynora Saxinger, Brent W. Footer, Emily G. McDonald, Todd C. Lee

## Abstract

**Importance:** Recent changes in the Infectious Diseases and Healthcare Epidemiology Societies of America (IDSA-SHEA) guidelines for managing *Clostridioides difficile* infections (CDI) have placed fidaxomicin as first-line treatment.

**Objective:** To estimate the net cost of first line fidaxomicin compared to vancomycin in the American and Canadian healthcare systems and to estimate the price points at which fidaxomicin would become cost saving. In Canada, net costs were from the public payer perspective. In the US, costs were from a healthcare and payer perspective.

**Data sources:** We identified all randomized controlled trials comparing fidaxomicin with vancomycin through the 2021 IDSA-SHEA guideline update. Medication costs were obtained from internet prices (US) and the Quebec drug formulary (Canada). The average cost of a CDI recurrence was established through a systematic review for each country.

**Study selection:** For fidaxomicin efficacy, we included double-blind and placebo-controlled trials. For the systematic review of recurrence costs, studies were included if they were primary research articles, had a cost-analysis of CDI, included cases of recurrent CDI, and were calculated with cost parameters from American or Canadian healthcare systems. Studies were excluded if the population was solely pediatric or hospitalized.

**Data extraction and Synthesis:** For the efficacy meta-analysis, data was pooled using a restricted maximal likelihood random effects model. For the cost review, the mean cost across identified studies was adjusted to reflect July 2022 dollars.

**Main Outcomes and Measures:** The primary outcome of the meta-analysis was CDI recurrence at Day 40. The primary outcome of the systematic review was the average cost of a CDI recurrence in the American and Canadian healthcare systems. The objective was to estimate the net cost per recurrence prevented and the price point below which fidaxomicin would be cost saving to either the public payer (Canada) or the insurer (US).

**Results:** The estimated mean system cost of a CDI recurrence was $15147USD and $8806CAD, respectively. At current drug pricing, to prevent one recurrence by using first line fidaxomicin over vancomycin would cost $43904USD (95%CI $35123-$65856) and $13,760CAD (95%CI $11,008-$20,640), respectively. The likelihood of fidaxomicin offering cost savings varies by country, with a 95% probability of fidaxomicin being cost saving if priced below $1180USD or $860CAD respectively.

**Conclusions and Relevance:** An increased drug expenditure on fidaxomicin will not be offset through recurrence prevention unless the fidaxomicin price is negotiated.

## Introduction

*Clostridioides difficile* infection (CDI) is a major cause of healthcare-associated diarrhea in North America. It is estimated that in 2017 there were nearly 462,000 cases in the United States (US) and in 2012^1^ there were approximately 37,900 cases in Canada^2^. Of these, 15-20% represent recurrences. The prevention of incident and recurrent episodes of CDI is therefore an important public health goal. Several pharmacologic and nonpharmacologic interventions have been investigated as initial treatment, and more specifically, to reduce risk of recurrence. For much of the twenty-first century, the recommended initial treatment of CDI has been oral metronidazole or vancomycin. In 2011, fidaxomicin was first demonstrated to be non-inferior to oral vancomycin for clinical cure^3^. This has ultimately been shown in 2 of 3 double-blind, randomized, placebo-controlled trials^3,4^, with all 3 providing evidence of a reduced risk of recurrence at day 40^3–5^. However, recommendations for fidaxomicin as first line therapy have lagged in guidelines and formulary uptake has been slow, presumably due to fidaxomicin’s higher cost. Issues surrounding affordability were highlighted in the 2017 Infectious Diseases and Healthcare Epidemiology Societies of America (IDSA-SHEA) guidelines^6^ and in the 2018 Association of Medical Microbiology and Infectious Diseases of Canada (AMMI) guidelines^7^. Now more than a decade since the initial trial was published, the 2021 update to the IDSA-SHEA *C. difficile* guidelines recommended fidaxomicin as first line therapy for all patients^8^. At the current pricing, treating all American and Canadian patients with fidaxomicin would cost an estimated $2.79 billion US dollars (USD) and $60 million Canadian dollars (CAD) per year, respectively. Whether the reduction of recurrent CDI will offset the higher up-front cost of fidaxomicin is unknown. We sought to estimate: 1) the net (added) cost of first line use of fidaxomicin required to prevent a recurrence as compared to oral vancomycin and compare this with 2) the cost of a CDI recurrence so that we could determine 3) the price point where a treatment course with fidaxomicin becomes cost saving.

## Methods

To estimate the comparative efficacy of fidaxomicin vs. vancomycin we conducted a meta-analysis of the three double blind, placebo-controlled, randomized controlled trials identified by IDSA-SHEA^8^ wherein fidaxomicin was compared head-to-head with vancomycin^3–5^. We excluded a fourth open-label trial which compared a longer total duration of fidaxomicin (30-days vs. 10-days in all other included studies)^9^. We examined the primary outcome of CDI recurrence at day 40, which was the longest common duration reported and meta-analyzed the risk ratio with 95% confidence intervals using a restricted maximum likelihood random effects model in STATA v. 17 (StataCorp LP). Using the overall control event rate as the expected baseline rate of recurrence, we then estimated the absolute risk difference, number needed to treat and associated 95% confidence intervals.

We obtained the lowest estimates for the American drug costs from GoodRx (fidaxomicin)^10^ and CostPlusDrugs (vancomycin)^11^. We obtained the Canadian drug costs from the Quebec formulary^12^ (the province with the highest rate of CDI). A 10-day course of fidaxomicin was estimated to cost $4452USD and $1,584 CAD, and that of vancomycin at $61USD (capsules) and $208 CAD (capsules). It is noted that some jurisdictions use the IV formulation as a PO treatment with consequent lower costs, but our comparison is based on commercial products. The difference in estimated costs and the NNT were used to calculate the additional cost per recurrence prevented.

We estimated the cost of a CDI recurrence in USD and CAD through a systematic review of the literature. Is the US, we assumed cost would be to an insurer and/or patient, and in Canada, to the public payer. We searched PubMed on July 10, 2022, with the search terms described in the Supplement. We included studies that were primary research articles, contained a cost-analysis of CDI, included cases of recurrent CDI, and were calculated with cost parameters based on the American or Canadian healthcare systems. Studies were excluded if the population studied was solely pediatric or hospitalized patients. References for all included studies were examined for additional applicable studies. Screening and data extraction was performed in duplicate (DP, JS, and TCL) with disagreement resolved by consensus. All costs were converted to May 2022 dollars using the Consumer Price Index Inflation Calculator^13^ (USD) and Bank of Canada Inflation Calculator^14^ (CAD) respectively. Across included studies, the average 2022-dollar cost was calculated and used for the analysis. We extracted the cost perspective that was examined in each of the included studies (e.g., public payer, traditional insurers, patient, societal, Medicare, Medicaid or third-party payer).

Finally, we calculated the probability of various effect sizes from the baseline recurrence rate and 95% confidence interval associated with the relative risk. We then identified how probable it was, at a specified price for fidaxomicin (rounded down to nearest $10), that the total cost of treating all patients with fidaxomicin relative to vancomycin would be offset by the cost savings from preventing recurrences (probability of cost equivalence). We created scatter plots of the probability of cost equivalence as a function of fidaxomicin price. For visualization purposes, a smooth line of best fit was generated with curvefit^15^ for STATA using a rational estimator.

## Results

### Fidaxomicin effectiveness

The overall relative risk for recurrence with 10 days of fidaxomicin vs. 10 days of vancomycin was 0.58 (95% CI 0.46-0.74; Figure 1). This corresponds to an absolute risk reduction of 10.8% (95% CI 6.7% - 14%) or a NNT of 10 (95% CI 8-15). At the current fidaxomicin and vancomycin prices, the estimated additional cost to prevent one recurrence in the USA was estimated as $43904USD (95%CI $35123-$65856) and in Canada this was estimated at $13,760CAD (95%CI $11,008-$20,640).

**Figure 1.**
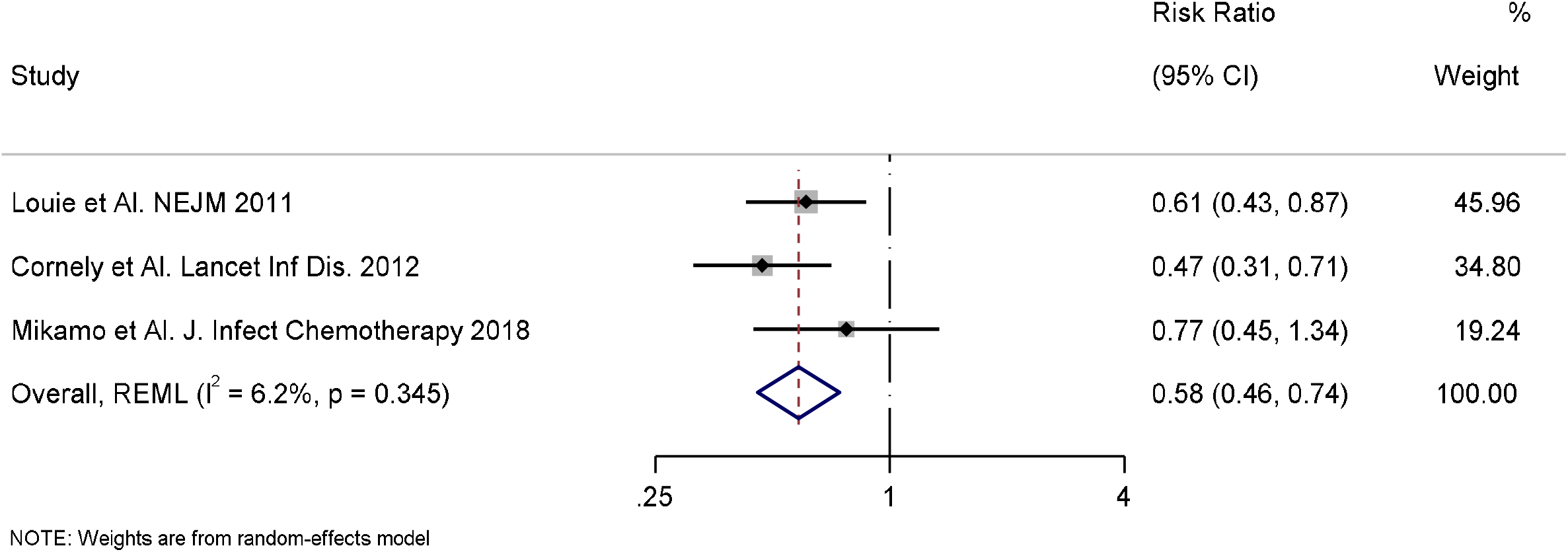
Forest Plot of Fidaxomicin Randomized Controlled Trials Risk of Recurrent CDI.

### Cost of recurrence

The results of the systematic literature review for the cost of a CDI recurrence in the American and Canadian healthcare systems are presented in Table 1. Additional descriptions of each included study are in the Supplement. For the USA, the initial search for the cost of a CDI recurrence yielded 786 results. Of these results, 110 articles were selected for further review. Of the 110 articles, 13 were reviews or meta-analyses, 36 included only hospitalized patients, 50 did not calculate the cost of a recurrent CDI episode, 3 included only a pediatric population, and 1 was based in the US. The 7 remaining articles were retained for the final analysis^16–22^. One article was subsequently excluded because it calculated the 12-month all-cause medical costs (as opposed to the attributable cost) of patients with recurrent CDI episodes^22^. Additionally, Luo et al. calculated the cost of recurrent CDI based on differing treatment strategies; the cost of the treatment with fidaxomicin was excluded from the overall average^16^.

**Table 1:** Summary of CDI recurrence cost by study

| Study | Recurrence Cost | May 2022 dollars | Cost perspectives |
| --- | --- | --- | --- |
| **United States** |  |  |  |
| McFarland et al.<sup>21</sup>, 1999 | \$1914 | \$3405.08 | Healthcare perspective; costs obtained from medical billing records and laboratory charges |
| Desai et al.<sup>18</sup>, 2016 | \$9501.74 | \$11722.81 | Healthcare perspective; study used societal perspective however indirect costs (productivity loss) were excluded from present analysis |
| Rodrigues et al.<sup>17</sup>, 2017 | \$34104 | \$41049.68 | Payer perspective; most cost values obtained from Centers for Medicare and Medicaid Services, with hospitalization cost from Healthcare Cost Utilization Project Nationwide Inpatient Sample (all-payer hospitalization database) |
| Zilberberg et al.<sup>20</sup>, 2017 | \$12043 | \$14495.70 | Payer perspective; costs measured as Medicare payments |
| Zhang et al.<sup>19</sup>, 2018 | \$10580 | \$12476.42 | Healthcare perspective; total healthcare costs were calculated as amount paid by primary and secondary insurers and by patients (i.e., copayment and deductibles) across all claims |
| Luo et al.<sup>16</sup>, 2020 | \$6826 | \$7734.25 | Modified third-party payer's perspective (included costs of medications, hospitalizations, and any procedures) |
| **Canada** |  |  |  |
| Wagner et al.<sup>24</sup>, 2014 | \$8250.05 | \$9961.71 | Public payer perspective |
| Levy et al.<sup>2</sup>, 2015 | \$8157.89 | \$9765.04 | Public payer perspective; study used a societal perspective however indirect costs were excluded from present analysis |
| Lapointe-Shaw et al.<sup>23</sup>, 2016 | Metronidazole:\$5386<br>Vancomycin:\$5929<br>Mean cost:\$5657.50 | Metronidazole:\$6351.97<br>Vancomycin:\$6692.35<br>Mean cost:\$6672.16 | Public payer perspective |

The search for the cost of a recurrence in Canada yielded 123 results, of which 18 articles were reviewed based on the title and abstract. Of these 18 studies, 14 studies were excluded: 5 studies did not include cases of recurrent CDI, 4 studies did not measure the cost of CDI, 4 studies were literature reviews, and 1 study measured the cost of readmission to hospital due to CDI without specifying if it was for first episode or recurrence. Four remaining studies included cases of recurrent or relapsed CDI and their cost^2,23–25^. One study that included cases of recurrent CDI was subsequently excluded as it presented the cost in median ($1812CAD), not mean values^25^. This left 3 studies that were included in the Canadian analysis for the cost of recurrence.

The estimated mean 2022 systemic costs for a recurrence of CDI in the American and Canadian healthcare systems respectively were $15147USD and $8806CAD. In the US, cost perspectives included payer and healthcare system perspectives, calculated using Medicare, third-party payers, and traditional insurers. In Canada, all studies reported costs based on a public payer perspective.

### Cost equivalence

With respect to the US, at the quoted price for 10 days of fidaxomicin and for 10 days of vancomycin capsules, there is a 0% chance that fidaxomicin will be cost equivalent by preventing the next CDI recurrence (Figure 2). At a price of approximately $1690 [$1630 more than the current cost of 10-day course of vancomycin] the probability of cost equivalence rises to 50% and at approximately $1180 [$1020 more than vancomycin] the probability rises to 95%. Therefore, fidaxomicin is very likely to be cost saving if priced below $1180 in the US. For Canada, at the current 10-day price of $1,580 CAD for fidaxomicin and $208 CAD for vancomycin, there is less than a 0.25% chance that fidaxomicin will be cost equivalent by preventing the next CDI recurrence (Figure 3). Reducing fidaxomicin price to approximately $1,150 CAD [$950 more than the current cost of a 10-day course of vancomycin] the probability of cost equivalence rises to 50% and at approximately $860 CAD [$660 more than vancomycin] the probability rises to 95%. In Canada, at any price below $860 CAD, fidaxomicin is likely to be cost saving.

**Figure 2.**
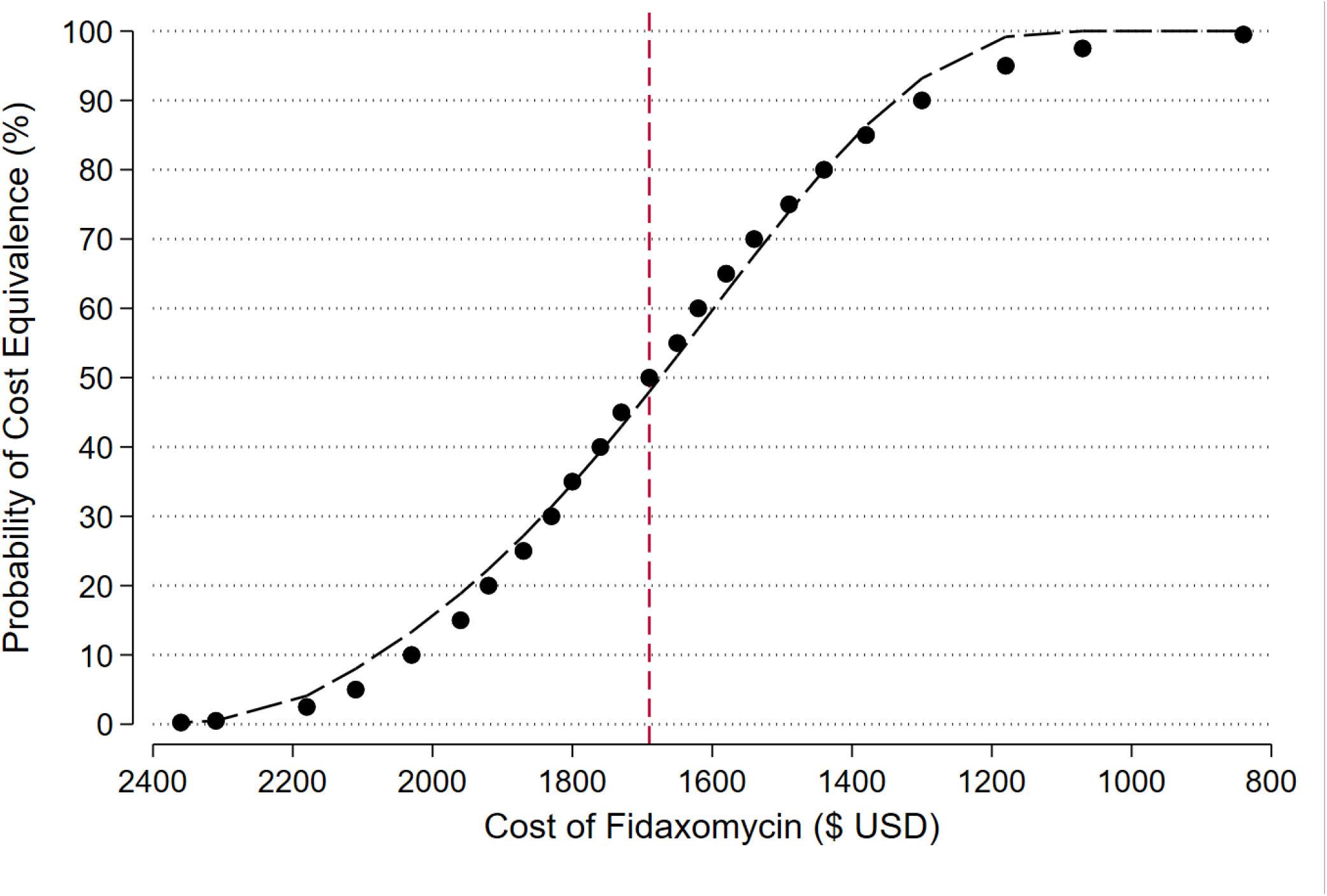
Probability of Fidaxomicin Cost Equivalence - USV.

**Figure 3.**
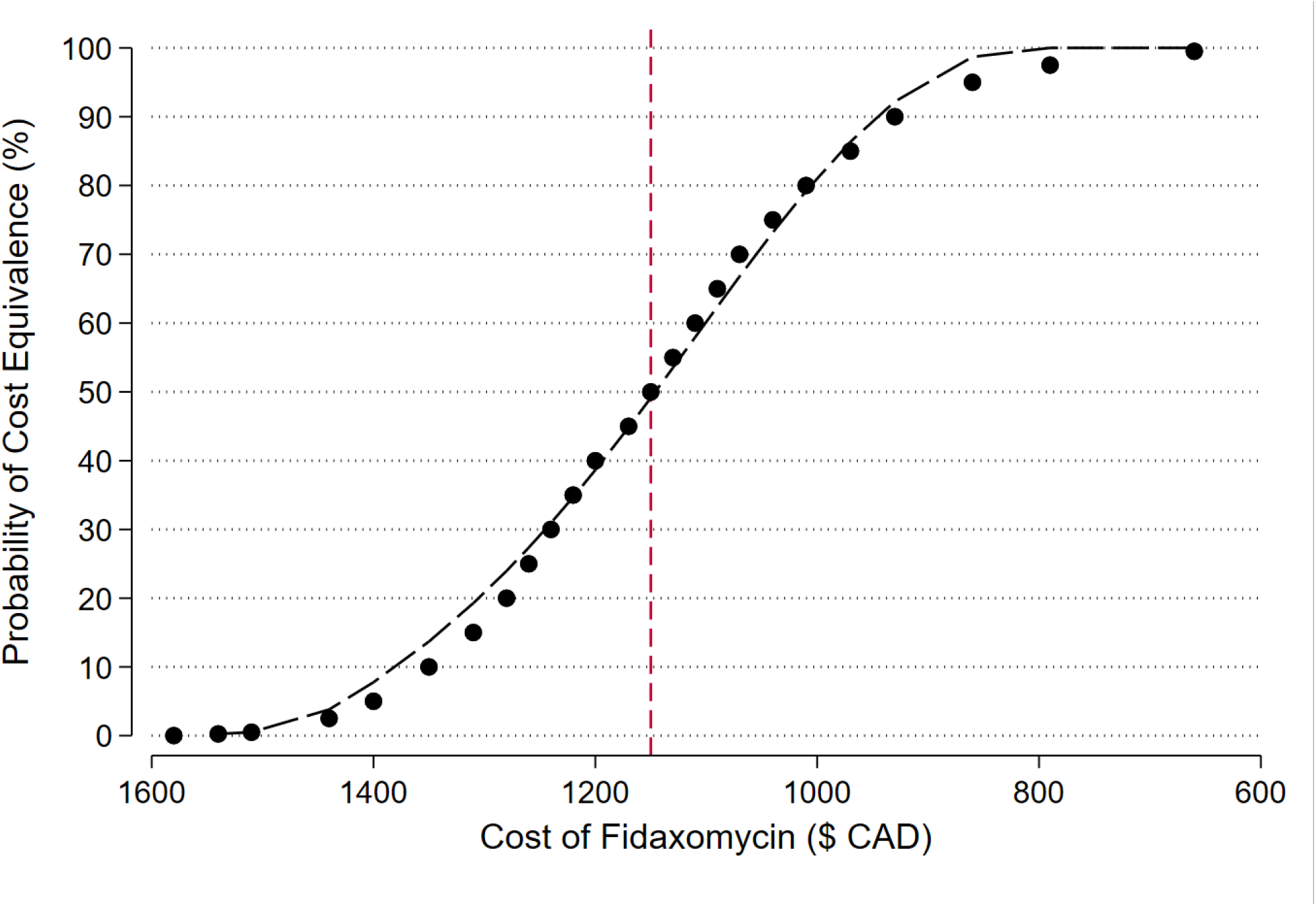
Probability of Fidaxomicin Cost Equivalence-Canada.

## Discussion

From our detailed review of the literature and associated calculations, we found that for both the US and Canada, the use of fidaxomicin as first-line treatment for CDI will cost substantially more to both the public payer in Canada and to US payers compared with potential cost savings through recurrence reduction. We identified price-points of approximately $1180 USD and $860 CAD at or below which the use of fidaxomicin is highly probable to be cost equivalent or cost saving. Despite double-blind randomized placebo-controlled trial evidence that 10 days of fidaxomicin is superior to 10 days of vancomycin for the secondary outcome of prevention of first recurrence at day 40, this efficacy has not translated into a meaningful uptake of fidaxomicin which we hypothesize is due to this very high financial impact. In Canada, individual provinces have their own drug plans, and negotiation with the manufacturer to obtain a more cost-equivalent price point is possible, which could facilitate a financially viable practice change. In the US, such negotiations are generally currently permitted by Medicare by law; however, negotiation of pricing could save the US billions per year for all drugs, including fidaxomicin^26^. Individual US insurance companies, particularly ones with large formulary budgets may have negotiating power to reduce costs.

This analysis has several limitations. At the current price of fidaxomicin, any strategy that increases the efficacy of vancomycin, for example, the use of an up-front decreasing dose taper to prevent recurrence (<u>NCT04138706</u>), would impact our results and would require recalculation. We have presented a best-case scenario for fidaxomicin by comparing it to a 10-day course of vancomycin. Furthermore, the efficacy of fidaxomicin to prevent recurrence at day 56 (the IDSA-SHEA definition of recurrence), or day 90, was not studied in the randomized trials. Up to 31% of recurrences may happen after day 42^27^ and there is no RCT data to allow comparisons including delayed recurrences. Fidaxomicin treatment outcomes have not been properly studied in patients with multiple recurrences, but it is possible that preventing the first recurrence could reduce the risk of subsequent recurrence and therefore be more attractive. Also, US drug prices are not fully transparent, and the costs borne to different parts of the system (patient, insurance company, hospitals/facilities) are often unclear. We used publicly available data to estimate the costs, but these costs may not reflect the costs to each party. Finally, further reducing the cost of vancomycin through the compounding of generic IV vancomycin into liquid form or reducing the cost of other formulations would increase the break-even price of fidaxomicin.

The estimation of CDI recurrence cost through systematic review for each country also has some limitations. The articles from the US had differing cost perspectives, with half the articles having a payer perspective while the other half had a healthcare perspective. The time frame of both Canadian and US studies also differed, ranging from 6 weeks of a recurrence to up to 12 months from a recurrence, with some studies having an unspecified time frame.

A strength of our study is the use of a meta-analytic assessment of the effect size for fidaxomicin from all the placebo-controlled trials, coupled with a systematic estimate of recurrence costs to produce a practical and easily understood comparison. Comparing additional drug costs vs. an estimate of the cost of a recurrence is a different analytic perspective that that cost per quality adjusted life-year point of view. Previous cost-effectiveness studies have been done, most showing a trivial fractional difference (e.g., 0.03) in quality-adjusted life-years (QALYs). More fundamentally, cost-effectiveness is not the same as cost saving. Cost-effectiveness measures, including cost per QALY and cost per incremental cost effectiveness ratio (ICER), assess added costs by a subjectively perceived threshold of value. Often this is contextualized against the historical price for a year of hemodialysis, which is lifesaving. However, hospitals, patients, and governments do not have unlimited budgets and most treatments are not a crucially lifesaving as hemodialysis. Even if an intervention is perceived as valuable, if the cost is unsustainable, cost-effectiveness may be irrelevant whereas cost-equivalence or cost-saving compared to current effective therapies are always relevant.

CDI causes a major burden to health systems worldwide and reduction of recurrence has value. Yet, health system sustainability requires thoughtful assessment of both current and future costs and benefits. At current pricing, a switch to first line fidaxomicin will cost billions of excess healthcare dollars to Canadian and US payers and based on this analysis these costs will not be recouped through the reduction of recurrent CDI. Assuming vancomycin costs remain the same, and until additional trials of novel vancomycin dosing strategies are available, a reduction of the cost of fidaxomicin to below $1180USD and $860CAD respectively would support a substantial change to fidaxomicin prescribing practices.

## Supporting information

CHEERS Checklist

## Data Availability

All data produced in the present study are available upon reasonable request to the authors

## Acknowledgements

TCL and EGM receive research salary support from the Fonds de Recherche du Québec – Santé.

## CRediT author statement

Conceptualization - TCL, DP; Methodology - TCL; Validation - TCL, DP; Formal Analysis - TCL, DP; Investigation - TCL, DP, JS; Resources - TCL; Data Curation - TCL, DP, JS; Writing - Original Draft - DP, EGM, TCL, JS; Writing - Review and Editing - All authors; Visualization - DP, TCL; Supervision - TCL; Project administration - TCL.

