## Supplement for "Fidaxomicin as first line: What will it cost in the USA and Canada?"

**Appendix 1:**

PubMed search terms:

Canadian analysis:
("Canada"[Mesh] OR "Canada" OR "Quebec") AND ("Clostridium Infections"[Mesh] OR "Clostridioides difficile"[Mesh] OR ("C. difficile" OR "Clostridium difficile" OR "Clostridioides difficile")) AND ("Costs and Cost Analysis"[Mesh] OR "Economics"[Mesh] OR "Cost-Benefit Analysis"[Mesh] OR "Health Care Costs"[Mesh] OR (cost OR costs OR economics))

USA analysis:
("United States"[Mesh] OR "United States" OR "USA") AND ("Clostridium Infections"[Mesh] OR "Clostridioides difficile"[Mesh] OR ("C. difficile" OR "Clostridium difficile" OR "Clostridioides difficile")) AND ("Costs and Cost Analysis"[Mesh] OR "Economics"[Mesh] OR "Cost-Benefit Analysis"[Mesh] OR "Health Care Costs"[Mesh] OR (cost OR costs OR economics))

### Appendix 2:

#### Table S1: Study summaries (Canada)

| Study | Population | Episode of CDI | Industry | Cost of CDI | Notes |
|---|---|---|---|---|---|
| Lapointe-Shaw 2016 | Model patient: 70-year-old community-dwelling person in Canada | Modeled for up to 3 recurrences<br><br>Does not include first episode of CDI | No industry funding | Fecal transplant by colonoscopy, enema, and NG tube: 5246, 5667, and 5935<br><br>Vancomycin: 5929<br>Metronidazole:5386<br>Fidaxomicin:7319 | Cost in 2014 Canadian dollars<br><br>Cost of fecal transplant not considered in this review as it is not a widely available option in Canada |
| Levy 2015 | Hospital, community, and long-term care infections in Canada | New and recurrent infections | Cubist Pharmaceuticals (manufacturer of fidaxomicin) | CDI-associated hospitalization, initial episode: 11930<br><br>CDI-associated hospitalization, recurrent episode:15330<br><br>Total cost of CDI relapses in 2012:<br>65100000<br><br>Total number of relapse cases in 2012: 7980<br><br>Cost per relapse (calculated):8157.89 | Cost in 2012 Canadian dollars<br><br>Number of hospital infections were estimated from mean estimated rate of persons infected in hospital per 10 000 bed-days multiplied by total patient days per province<br><br>Number of community and long-term care infections were estimated by using the known ratio of hospital to community infections in Manitoba and applied to each province<br><br>Number of recurrences were calculated by estimated proportion of CDI that are recurrent (0.271). Recurrences included relapses and reinfections. |
| Wagner 2014 | Patients with severe CDI as defined by clinical guidelines | Subgroup analysis of patients with recurrent CDI | No industry funding | Cost of treatment for 1000 patients with vancomycin of first recurrence:8250046<br><br>Cost per recurrence (calculated):8250.05 | Model input parameters for costs in this study ranged from 2010-2013, with the hospitalization cost in 2010 CAD: cost conversion from assumed currency of 2010 Canadian dollars<br><br>This cost assumes a hospitalization rate of 63.8% for recurrences of CDI |

**Table S2: Study summaries (USA)**

| Study | Population | Episode of CDI | Industry | Cost of CDI | Notes |
|---|---|---|---|---|---|
| Luo 2020 | Model patient: 65-year-old community dwelling patient | First recurrence of mild to moderate CDI | No industry funding | Based on treatment:<br>Vancomycin:7006<br>Fidaxomicin:7557<br>Bezlotoxumab:9612<br><br>FMT via colonoscopy, capsules: 5250 and 5436<br><br>Overall average of all treatments excluding fidaxomicin:6826 | 2019 USD |
| Rodrigues 2017 | Adult patients | Recurrent CDI | Funding from Cubist/Merck (manufactures fidaxomicin) | 34104 | 2016 USD; Costs associated with CDI recurrences for up to a year after recurrent episode |
| Desai 2016 | Adult and pediatric populations | Recurrent CDI in 4 settings: hospitals, long-term care, long-term acute care hospitals, community | Funding from Merck (manufactures fidaxomicin) | Total healthcare cost of recurrent CDI:1,585,089,451<br>Total cases of recurrent CDI:166,821<br>Cost per recurrence (calculated):9501.74 | 2014 USD |
| Zhang 2018 | Adult and pediatric patients | Recurrent CDI | Funding from Merck (manufactures fidaxomicin) | Average attributable healthcare cost of an episode of recurrent CDI for 6 months post:10580 | 2014 USD |
| Zilberberg 2017 | Patients aged >=65 years, in nursing homes | CDI recurrences | Funding from Merck (manufactures fidaxomicin) | Excess costs associated with recurrence of CDI:12043 | Currency assumed to be 2017 USD |
| McFarland 1999 | Adult patients | Recurrent CDI | Funding from Laboratoires Biocodex (manufactures probiotics) | Cost of recurrent CDI:1914 | Currency assumed to be 1999 USD |
